# Commercial immunoglobulin products now contain neutralising antibodies against SARS-CoV-2 spike protein which are detectable in patient serum

**DOI:** 10.1101/2022.09.22.22280216

**Authors:** Vinit Upasani, Mary O’Sullivan, Fernando Moreira, Sarita Workman, Andrew Symes, Siobhan O Burns, Susan Tadros, Laura McCoy, David M Lowe

**Affiliations:** Institute of Immunity and Transplantation, University College London, London, UK; Department of Clinical Immunology, Royal Free London NHS Foundation Trust, London, UK

**Keywords:** SARS-CoV-2, spike antibody, IVIG, neutralisation, immunodeficiency

## Abstract

Antibody-deficient patients respond poorly to COVID-19 vaccination and are at risk of severe or prolonged infection. Prophylaxis with anti-SARS-CoV-2 monoclonal antibodies has been considered. We here demonstrate that many immunoglobulin preparations now contain neutralising anti-SARS-CoV-2 antibodies which are transmitted to patients in good concentrations, albeit with significant differences between products.

## Text

Primary and secondary antibody deficiencies are characterised by impaired ability to mount a functional humoral immune response (1). Patients with these conditions have either significantly decreased or no antibody responses after most infections or vaccinations compared to healthy individuals (1), including to SARS-CoV-2 (2,3). These patients rely on long-term infusions with commercial intravenous immunoglobulin (IVIG) products, which is collectively referred to as Immunoglobulin Replacement Therapy (IRT), for protection against infectious diseases. The IVIG products used for IRT are derived from the plasma of thousands of pre-screened healthydonors containing natural antibodies against various antigens.

Since the beginning of the SARS-CoV-2 pandemic, individuals with antibody deficiencies have been at considerable risk from the infection and have shown increased hospitalisation rates and mortality (2,4). They are also at risk of prolonged and relapsing COVID-19 disease (3). In the absence of a robust humoral vaccination response, attention has been given recently to other potential prophylactic strategies such as long-acting monoclonal antibodies against the spike (S) protein of SARS-CoV-2 (5). We hypothesised that, given the ubiquity of vaccination and/or natural infection even by 2021, immunoglobulin preparations prepared from plasma of healthy donors may now contain levels of antibody that confer some protection, reducing the imperative for other prophylaxis. Therefore, in this study, we evaluated the antibody response against SARS-CoV-2 in a cohort of patients before and after undergoing infusion with commercial IVIG products. Furthermore, in these paired samples and the products themselves, we estimated the neutralising antibody titres against SARS-CoV-2 wildtype virus and Omicron variant using an *in vitro* luminescence-based neutralisation assay.

We recruited 35 individuals with immunodeficiencies on IRT and 7 healthy controls. All donors provided written informed consent under protocols approved by a UK National Health Service (NHS) research ethics committee (Hampstead Research Ethics Committee, Refs 04/Q0501/119 and 08/H0720/46). Blood sampling was performed from the patients in the clinic before and immediately after an IVIG infusion was completed. The patient demographic data are summarised in Supplementary Table 1. The most common diagnoses were common variable immunodeficiency (CVID) and secondary hypogammaglobulinemia. All patients had been on regular IVIG for at least 6 months and IgG trough levels were generally >7 g/L. Patients at our centre receive a range of immunoglobulin products with random selection, unless there are specific contraindications or adverse reactions. All patients had received at least 2 vaccinations against SARS-CoV-2. 20 patients reported a prior history of COVID-19 (two patients reported 2 infections) with around half of episodes treated and others usually resolving spontaneously. Of note, one patient had received sotrovimab treatment within the previous 2 weeks.

First, we determined the titres of antibodies directed against SARS-CoV-2 spike protein using a commercial immunoassay (Elecsys® Anti-SARS-CoV-2 S, Roche). Samples with titres >2500 U/mL were diluted 10-fold and repeated, resulting in a maximum titre of 25,000 U/mL. Most patients showed a significant increase in anti-S antibody titres post-infusion with immunoglobulin products (Figure 1A) (p<0.01) with the median titre increasing from 2123 U/mL pre-infusion to 10600 U/mL post-infusion. Upon classification based on IVIG product administered, this increase in anti-S antibody titres was observable in most patients receiving Privigen or Octagam. Responses in patients receiving Intratect were variable, perhaps indicating batch to batch variation. Post-infusion titres in patients who received Flebogamma were relatively modest while for Iqymune, S antibody titres remained unchanged or decreased slightly and some were very low (<100 U/mL). The patient who had received sotrovimab recently had a titre of >25,000 U/mL even pre-infusion (receiving Privigen). Three patients receiving Intratect had similarly high titres pre-infusion even in the absence of recent treatment.

**Figure 1:**
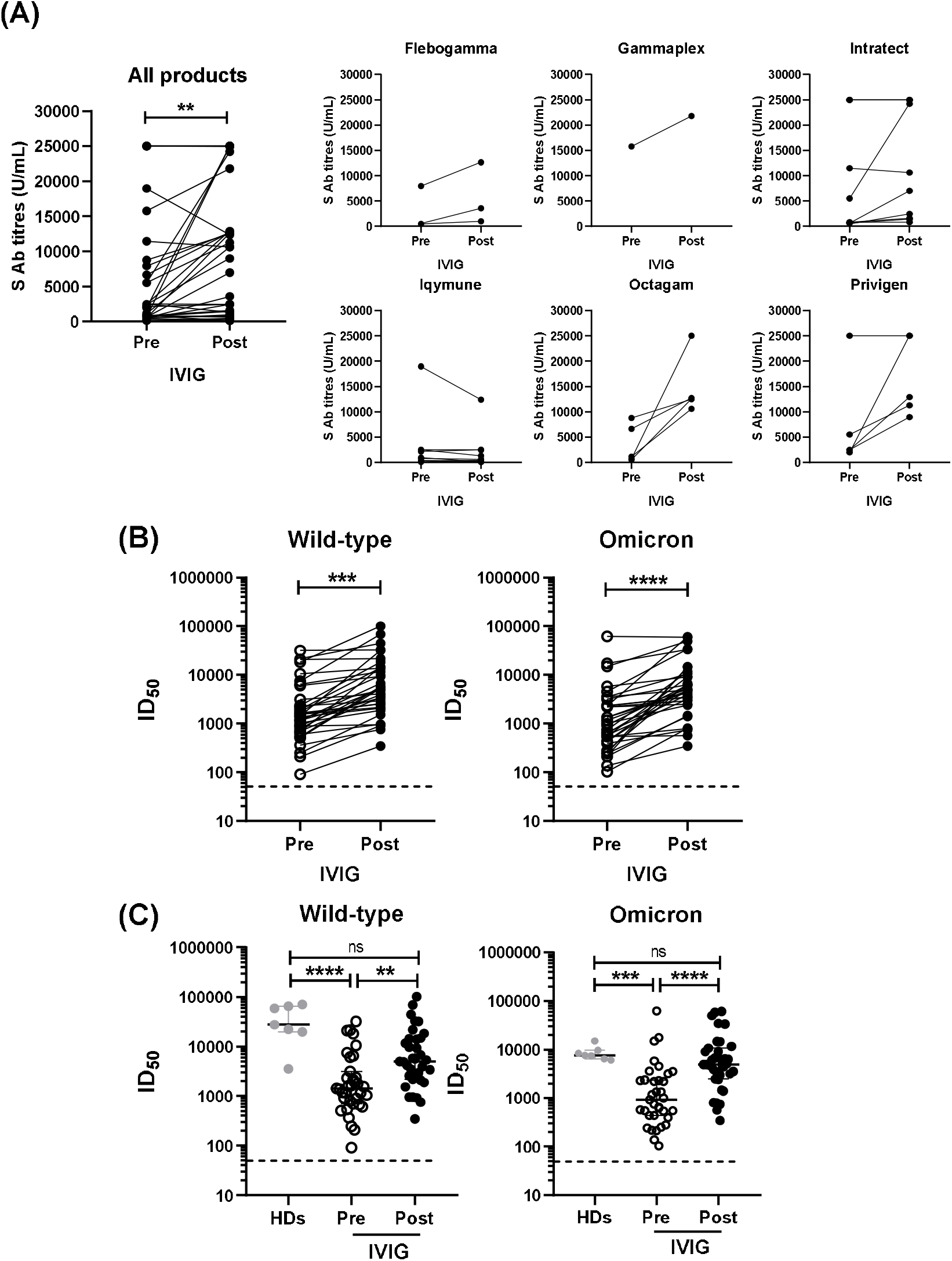
Humoral immune responses in patients with PIDs receiving IVIG products. **A**. SARS-CoV-2 S Ab titres in serum samples taken from patients pre- and post-infusion with IVIG products: results are presented from all patients and according to product received. **B**. Comparison of neutralising antibody titres (ID_50_) against SARS-CoV-2 wild-type and Omicron viruses in serum samples from patients pre- and post-infusion with IVIG products. **C**. Neutralising antibody titres in comparison with healthy donors. *P*-values were calculated using Wilcoxon paired t test for comparing two groups and Kruskal-Wallis test with Dunn’s post-hoc test for comparing three groups (\*\**P*<0.01; *** *P*<0.001; **** *P*<0.0001).

Next, neutralising antibody titres against wild-type SARS-CoV-2 and Omicron were determined using a luminescence-based neutralisation assay as described previously (6). Briefly, serially diluted serum samples or neat immunoglobulin products were incubated in a 96-well plate with a HIV-based pseudovirus expressing S protein of wild-type SARS-CoV-2 virus or the omicron variant on its surface. HeLa cells engineered to express ACE2, a surface attachment factor for SARS-CoV-2 (7), were added to the respective wells. After 3 days of incubation, cells were lysed, and plates were read upon addition of a luminescent substrate. Neutralisation capacity of the serum sample is expressed in terms of inhibitory dilution 50 (ID_50_), i.e., the serum dilution at which 50% of infection is inhibited compared to the virus alone. Neutralisation titres against both wild-type and Omicron variants significantly increased post-infusion with IVIG products (Figure 1B). However, baseline neutralisation titres against wild-type virus were higher than Omicron prior to infusion with IVIG products (median ID_50_ : 1437 vs 925).

We also compared the SARS-CoV-2 neutralisation titres of patients with immunodeficiencies undergoing IVIG infusions to healthy individuals. Neutralising antibody titres against both wild-type and Omicron viruses were significantly lower in patients prior to infusion with IVIG products compared to healthy donors (median ID_50_: Wild-type 1437 vs 27574; Omicron 925 vs 7563, p<0.0001 and p<0.001 respectively) but were restored to levels comparable to those observed in healthy donors after infusion, albeit with considerable heterogeneity (median ID_50_: Wild-type 4985 vs 27574; Omicron 4932 vs 7563, no statistically significant differences) (Figure 1C). There were no statistical differences between S Ab concentrations or neutralisation titres (pre- or post-infusion) between patients with a history of COVID-19 in the last 6 months versus those who had COVID-19 more than 6 months previously.

To confirm that neutralisation was derived from the immunoglobulin products, we tested some of the corresponding batches of IVIG products (except Gammaplex) used on those days in the clinic at neat concentration followed by 5-fold serial dilutions in the SARS-CoV-2 neutralisation assay. All products demonstrated detectable neutralisation varying between ID50 of 1:25000 and 1:2×10^7^, higher titres overall than observed in patient serum. However, in general products showed lower neutralisation activity against Omicron compared to the wild-type virus (Supplementary Figure 1A) in line with observations in convalescent and vaccinated populations. Batches of Intratect and Privigen showed significantly higher neutralisation activity against wild-type virus compared to other products, especially Flebogamma and Iqymune, which is in line with S Ab results observed previously. Octagam neutralised the Omicron variant poorly compared to wild-type virus, while Iqymune neutralised Omicron somewhat better than wild- type virus despite patients on Iqymune generally having lower anti-S and neutralising antibody titres (Supplementary Figure 1A). However, maximum titre against either virus with this product was below 1:10^5^.

To ascertain whether these differences between IVIG products are reflected in the neutralising antibody titres of patient serum, post-infusion neutralisation titres were compared based on the IVIG product received. Lower neutralisation against wild-type virus was seen in serum from patients who received Iqymune, consistent with other results, but there were no clear differences between products for Omicron. However, a high degree of patient-dependent variability in neutralisation titres was observed in our cohort (Supplementary Figure 1B). This is likely to reflect varying immunological outcomes from prior vaccination or infection with some patients able to generate an antibody response and others not. However, the overall improvement in neutralisation across the cohort from pre- to post-infusion (Figure 1) suggests that most IVIG products are conferring additional benefit, even against omicron variants. We have not here investigated subcutaneous immunoglobulin products although we anticipate that findings would be similar.

In summary, we have demonstrated that, in mid-2022, most commercial IVIG preparations now contain neutralising anti-SARS-CoV-2 spike antibodies and that these are detectable in patient serum after infusion. The majority of people have good levels of S antibody, at levels that would render them ineligible for current prophylaxis studies (eg https://clinicaltrials.gov/ct2/show/NCT04870333). Although antibody-deficient patients have lower neutralisation capacity than healthy controls immediately pre-infusion (at IgG trough), this is largely restored by infusion. However, there is considerable variability between products and potentially between batches: this presumably relates to the timing of plasma harvest and is likely to improve over time. The impact of this heterogeneity on neutralisation of omicron variants is less clear and we note that many of our patients had previously had COVID-19, albeit with full clinical recovery. However, this finding should reassure patients and clinicians that they are likely to have at least relative protection against COVID-19.

## Data Availability

All data produced in the present work are contained in the manuscript.

## Acknowledgements

The authors would like to thank Rosemarie Ford and Dylan Jankovic of Institute of Immunity and Transplantation, UCL for their technical input with the neutralisation assay.

## Supplementary Material

**Supplementary table 1:** Patient demographics and clinical details. CVID – common variable immunodeficiency; XLA – X-linked agammaglobulinemia; IVIG – intravenous immunoglobulin; g/L – grams per litre

|  |  |  |
| --- | --- | --- |
| Age (years) | Median<br>Range | 60<br>20 – 86 |
| Sex | Male<br>Female | 12<br>23 |
| Diagnosis | CVID<br>XLA<br>Other / undefined primary<br>hypogammaglobulinemia<br>Secondary hypogammaglobulinemia | 12<br>2<br>9<br>12 |
| Duration on IVIG | 6 months – 1 year<br>1 – 3 years<br>>3 years | 1<br>4<br>30 |
| Infusion frequency | 2-weekly<br>3-weekly<br>4-weekly<br>5-weekly<br>6-weekly | 1<br>4<br>18<br>3<br>9 |
| Last IgG trough<br>(g/L) | Median<br>Range | 9.3<br>4.1 – 13.7 |
| Received COVID-19 vaccination | No<br>Yes<br>2 vaccinations<br>3 vaccinations<br>4 vaccinations<br>5 vaccinations | 0<br>35<br>3<br>7<br>19<br>6 |
| Time since last<br>vaccination (days) | Median<br>Range | 124<br>27 – 410 |
| Known previous<br>COVID-19 | No<br>Yes, once<br>Yes, twice | 15<br>18<br>2 |
| COVID-19 episodes | Untreated<br>Sotrovimab treatment<br>Remdesivir treatment<br>Nirmatrelvir/ritonavir treatment<br>Unknown treatment | 10<br>5<br>1<br>1<br>5 |
| Time since COVID-19 (days) | Median<br>Range | 131<br>9 – 857 |

**Supplementary figure 1:**
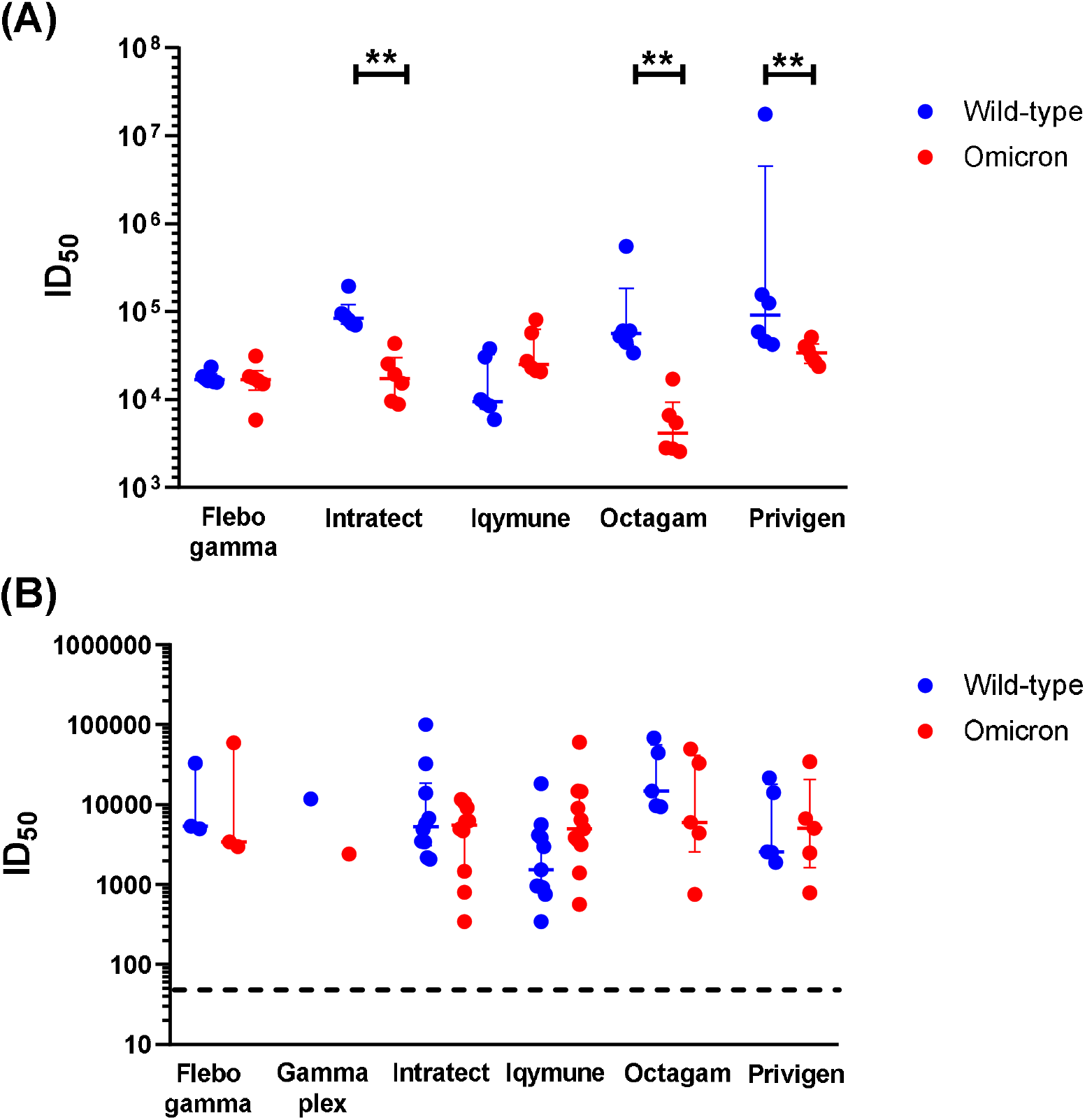
**(A)** Product wise comparison of neutralising antibody titres (ID_50_) against SARS-CoV-2 wild-type and Omicron viruses in neat IVIG products used for infusions in patients. **(B)** Neutralising antibody titres (ID_50_) against SARS-CoV-2 wild-type and Omicron viruses in patient serum post-infusion with IVIG products. *P*-values were calculated using non-parametric Mann-Whitney test (\**P*<0.05; \*\**P*<0.01).

